# Trajectory of viral load in a prospective population-based cohort with incident SARS-CoV-2 G614 infection

**DOI:** 10.1101/2021.08.27.21262754

**Authors:** Helen C. Stankiewicz Karita, Tracy Q. Dong, Christine Johnston, Kathleen M. Neuzil, Michael K. Paasche-Orlow, Patricia J. Kissinger, Anna Bershteyn, Lorna E. Thorpe, Meagan Deming, Angelica Kottkamp, Miriam Laufer, Raphael J. Landovitz, Alfred Luk, Risa Hoffman, Pavitra Roychoudhury, Craig A. Magaret, Alexander L. Grenninger, Meei-Li Huang, Keith R. Jerome, Mark Wener, Connie Celum, Helen Y. Chu, Jared M. Baeten, Anna Wald, Ruanne V. Barnabas, Elizabeth R. Brown, the Hydroxychloroquine COVID-19 PEP Study Team

## Abstract

**Importance:** SARS-CoV-2 viral trajectory has not been well-characterized in documented incident infections. These data will inform SARS-CoV-2 natural history, transmission dynamics, prevention practices, and therapeutic development.

**Objective:** To prospectively characterize early SARS-CoV-2 viral shedding in persons with incident infection.

**Design:** Prospective cohort study.

**Setting:** Secondary data analysis from a multicenter study in the U.S.

**Participants:** The samples derived from a randomized controlled trial of 829 community-based asymptomatic participants recently exposed (<96 hours) to persons with SARS-CoV-2. Participants collected daily mid-turbinate swabs for SARS-CoV-2 detection by polymerase-chain-reaction and symptom diaries for 14-days. Persons with negative swab for SARS-CoV-2 at baseline who developed infection during the study were included in the analysis.

**Exposure:** Laboratory-confirmed SARS-CoV-2 infection.

**Main outcomes and measures:** The observed SARS-CoV-2 viral shedding characteristics were summarized and shedding trajectories were examined using a piece-wise linear mixed-effects modeling. Whole viral genome sequencing was performed on samples with cycle threshold (Ct)<34.

**Results:** Ninety-seven persons (57% women, median age 37-years) developed incident infections during 14-days of follow-up. Two-hundred fifteen sequenced samples were assigned to 15 lineages that belonged to the G614 variant. Forty-two (43%), 18(19%), and 31(32%) participants had viral shedding for 1 day, 2-6 days, and ≥7 days, with median peak viral load Ct of 38.5, 36.7, and 18.3, respectively. Six (6%) participants had 1–6 days of observed viral shedding with censored duration. The peak average viral load was observed on day 3 of viral shedding. The average Ct value was lower, indicating higher viral load, in persons reporting COVID-19 symptoms than asymptomatic. Using the statistical model, the median time from shedding onset to peak viral load was 1.4 days followed by a median of 9.7 days before clearance.

**Conclusions and Relevance:** Incident SARS-CoV-2 G614 infection resulted in a rapid viral load peak followed by slower decay and positive correlation between peak viral load and shedding duration; duration of shedding was heterogeneous. This longitudinal evaluation of the SARS-CoV-2 G614 variant with frequent molecular testing may serve as a reference for comparing emergent viral lineages to inform clinical trial designs and public health strategies to contain the spread of the virus.

**KEY POINTS:** *Question:* What are the early SARS-CoV-2 G614 viral shedding characteristics in persons with incident infection?

*Findings:* In this prospective cohort of 97 community-based participants who collected daily mid-turbinate swabs for SARS-CoV-2 detection after recent exposure to SARS-CoV-2, viral trajectory was characterized by a rapid peak followed by slower decay. Peak viral load correlated positively with symptoms. The duration of shedding was heterogeneous.

*Meaning:* A detailed description of the SARS-CoV-2 G614 viral shedding trajectory serves as baseline for comparison to new viral variants of concern and inform models for the planning of clinical trials and transmission dynamics to end this pandemic.

## Introduction

Since December 2019 more than 4 million people worldwide have died from COVID-19, caused by SARS-CoV-2. By late 2020, highly efficacious vaccines became available under emergency authorization use (EUA) in several countries.^1–4^ Despite the substantial advancement in public health prevention approaches and deployment of vaccines for curbing the pandemic, the viral spread continues as less than 30% of the global population have received any vaccine doses.^5^ Furthermore, the emergence and spread of novel variants have compelled global attention because of increased transmissibility and potentially virulence and have threatened the control of the pandemic.^6–10^ Novel variants, including the newly described delta (Pango B.1.617.2 lineage), are outcompeting the wild-type virus and the earlier-identified variants such as alpha variant (Pango B.1.1.7 lineage) as the dominant circulating strain in U.S. and many other countries.^6,8,11,12^ Preliminary evidence suggests a reduction in neutralizing activity of some antibodies in convalescent plasma and vaccines against strains with reports of reduced efficacy against prevention of infection for existing vaccines.^13–17^

Substantial uncertainty regarding SARS-CoV-2 natural history and early viral shedding remains. Studies of frequently monitored hospitalized patients with COVID-19 have provided data on SARS-CoV-2 nasal detection characteristics from diagnosis through the course of the infection.^18–21^ Among outpatients, very few prospective studies have captured viral dynamics and symptoms from the acquisition of infection rather than the time of diagnosis.^22,23^ Furthermore, little data exist on early viral shedding after infection with new variants of concern and their differences compared to the SARS-CoV-2 Wuhan strain carrying the D614G Spike mutation (G614 virus) identified in early cases.

Emerging reports of SARS-CoV-2 variants of concern (VOC) have suggested a higher viral load in respiratory samples and more prolonged shedding than early variants.^24–27^ As SARS-CoV-2 continued spread allows for the appearance of new viral mutations and potentially deleterious variants, the characterization of early viral shedding trajectories is essential to inform the likelihood of transmission and to improve models of transmission dynamics to optimize appropriate infection control interventions. With the threat of emerging resistance to passive immunity from monoclonal antibodies and acquired immunity generated by current COVID-19 vaccines, frequent viral load measurements in clinical trials evaluating new therapeutics are crucial to measure objective disease markers and allow accurate assessment of therapy impact.

Prior to the documented emergence of new SARS-CoV-2 VOC, we enrolled asymptomatic persons exposed to SARS-CoV-2 in a prospective study. Participants self-collected mid-turbinate swabs daily for 14-days within a median of 1-2 days after exposure to household, patient, or social contacts with virologically documented SARS-CoV-2. We examined the SARS-CoV-2 viral shedding using real-time reverse-transcription polymerase chain reaction (RT-PCR) assay in participants with a negative swab at baseline and evaluated lineage assignments by whole genome sequencing. The results provide detailed SARS-CoV-2 G614 strain trajectories to establish a baseline for comparison to new viral variants detected in genomic surveillance programs.

## Methods

### Study population and procedures

The population was a subset of the Hydroxychloroquine (HCQ) COVID-19 PEP Study, a double-blinded, household-randomized controlled trial comparing HCQ to placebo-like control for prevention of SARS-CoV-2 infection.^28^ The study was conducted between March and August 2020 and enrolled 829 asymptomatic close contacts (defined as household residing in the same residence of an index case, prolonged exposure in a confined space, and/or healthcare worker who cared for index case without appropriate personal protective equipment) recently exposed (<96 hours) to persons with laboratory-confirmed SARS-CoV-2 infection. The study found no differences between HCQ and placebo-like control in SARS-CoV-2 acquisition, and no participants received EUA COVID-19 vaccines. As such, the trial allowed an examination of the natural history of SARS-CoV-2 among participants regardless of assigned group. For the present analysis, we evaluated the subset of participants with a negative swab for SARS-CoV-2 at baseline who acquired SARS-CoV-2 during the study follow-up. The Western Institutional Review Board approved the trial; all participants provided informed consent.

Participants self-collected daily mid-turbinate swabs for 14 consecutive days. The participant placed the swab in a collection tube containing 1mL of phosphate-buffered saline, and such specimens were shipped to the University of Washington (UW) every 5-7 days.^28^ Participants completed daily symptom questionnaires using REDCap.^29^

### Laboratory methods

Mid-turbinate swabs were tested for SARS-CoV-2 RNA at the UW Virology Laboratory using RT-PCR targeting the SARS-CoV-2 nucleocapsid genes N1 and N2 as described previously.^30^ To document adequate swab technique as indicated by the presence of cellular DNA, a subset of swabs (n=176, 13% of collected swabs from 37 participants) were tested for the human DNA maker RNase P (42 swabs were SARS-CoV2 positive and 134 negative).

Whole viral genome sequencing was attempted on all samples that had a Cycle threshold (Ct)<34. Sequencing libraries were prepared using the Swift Biosciences’ Normalase amplicon panel^31^ and sequenced on Illumina Nextseq instruments using 2×150 reads. Genomes were assembled using a custom pipeline described previously.^31^ Genomes with <10% Ns were selected for further analysis including clade assignment which was performed using the Pangolin (https://pangolin.cog-uk.io/, 10.1038/s41564-020-0770-5) and Nextclade (https://clades.nextstrain.org/) tools.

### Statistical analysis

#### Observed viral shedding characteristics

Viral shedding duration was defined as the number of days from the first positive self-collected mid-turbinate swab until the last positive swab. Censoring was indicated if a participant had a positive swab on the last day of swab collection. For a given day, a participant’s viral load was calculated as the average of N1 and N2 Ct values from the RT-PCR test. The Ct value assigned to the negative samples was 40. The cohort was divided into four shedding duration sub-groups based on interval testing in prior longitudinal studies of viral trajectories^32,33^: 1 day, 2-6 days, 7 days or more, and censored with 1-6 days of observed shedding. Participants reached the observed peak viral load when the swab collected yielded the minimum average Ct value indicating the highest quantity of virus detected. The Kaplan-Meier method was used to calculate the median peak viral load in Ct and the associated 95% confidence interval (CI) within each duration sub-group, accounting for censoring and clustering at the household level. The individual observed peak viral load in Ct and the medians were plotted within each shedding duration sub-group.

The average SARS-CoV-2 viral load by day of viral shedding was plotted among all participants with incident infection, within those who reported COVID-19 symptoms^34^ and those who remained asymptomatic throughout the study follow-up. The time and magnitude of the average peak viral load were reported and compared across the groups.

#### Model-based viral shedding characteristics

Characteristics of viral shedding trajectories were estimated using a piece-wise linear mixed-effects model (see Appendix for details). To fit the model, we used data from all participants who had 2 or more days of viral detection in the parent study, including participants with a positive swab at baseline. The model estimated the average rate of increase and decrease in viral load, the duration of shedding, and the peak viral load for each participant, which are presented graphically. The population-level viral shedding trajectory estimates were conditional on the duration of shedding and were summarized by grouping participants based on the model-based duration and plotting the mean viral load by day since onset within each group.

## Results

### Study participants

Among 829 participants enrolled in the parent study, 97 (12%) had a negative SARS-CoV-2 swab at baseline and at least one subsequent positive swab during the 14-days of follow-up. In the cohort with incident infection, 55 participants (57%) were women and 42 (43%) men. Participant median age was 37 years (interquartile range, IQR= 27 – 52 years). Participants identified as non-Hispanic White (n=53, 55%), Asian (n=17, 18%), African American (n=10, 10%), or American Indian/Alaska Native (n=2, 2%). Twenty-nine (30%) participants identified as Hispanic.

We obtained 1332 swabs from 97 participants, of whom 1 (1%) provided 1-6 swabs, 6 (6%) provided 7-13, and 90 (93%) provided 14. Of these, 378 (28%) swabs were positive for SARS-CoV-2 and 954 (72%) were negative. The median time to first mid-turbinate swab collection was 43 hours (IQR=21 – 74) since the last exposure to index case(s). RNase P was detected in all the specimens tested (**Figure S6 in the Supplement**).

A total of 215 samples with Ct<34, corresponding to 42 participants were sequenced. The sequenced genomes were assigned to 15 lineages that belonged to the G614 variant (**Table S1 in the Supplement**).

### Observed SARS-CoV-2 characteristics among participants with incident infection

Individual viral load trajectories for the 97 persons with incident SARS-CoV-2 infection are presented in **Figure S4 in the Supplement**. Seventy-three (75%) participants were assumed to have a completely observed shedding duration (i.e., first and last swabs were negative) and 24 (25%) had censored shedding (i.e., the last swab was positive). Of 97 participants, 42 (43%) had viral detection for 1 day, 18 (19%) for 2-6 days, 31 (32%) for 7 or more days (**Figure 1**). Six (6%) persons had 1-6 days of observed shedding but were right-censored. A lower median peak Ct (indicating higher levels of viral RNA) was observed in participants with longer shedding duration. The median peak Ct viral load was 38.5 (95% CI, 38.33 - 39.0), 36.7 (95% CI, 30.2 - 38.1), and 18.3 (95% CI, 17.4 - 22.0) in persons with viral shedding of 1 day, 2-6 days, and 7 or more days, respectively (**Figure 1**).

**Figure 1.**
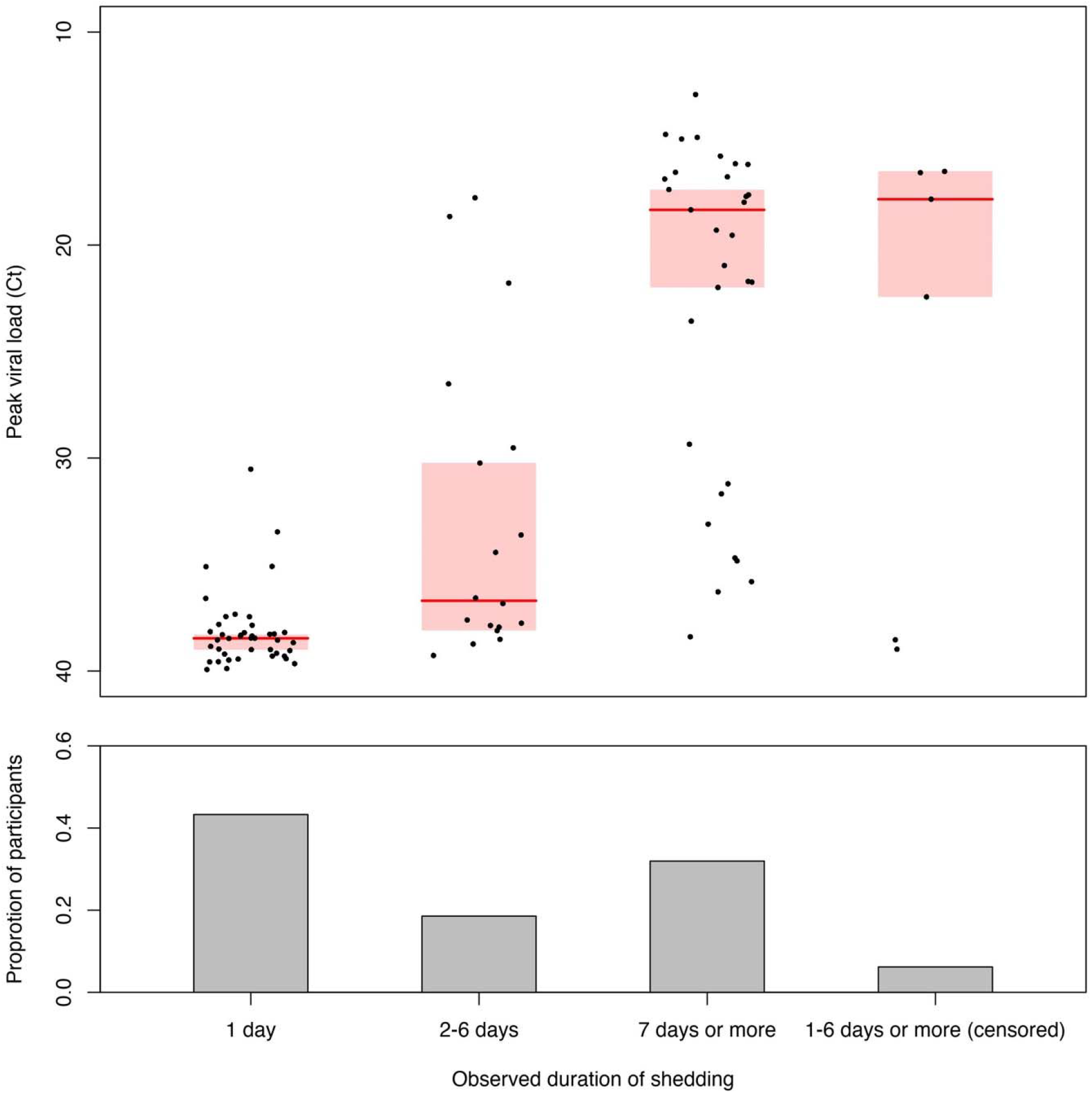
The observed shedding duration and peak viral load among 97 participants with incident Severe Acute Respiratory Syndrome Coronavirus-2 (SARS-CoV-2) infection. We grouped the participants according to the observed duration of shedding in days. Each dot represents a participant’s observed peak viral load, defined as the minimum average cycle threshold (Ct) value measured during the study follow-up. The horizontal line segments indicate the median peak viral load in each shedding duration group, and the shaded areas indicate the 95% confidence intervals (CI) around the medians, calculated using Kaplan-Meier method that accounted for censoring and clustering within households. The vertical bars in the lower panel represent the proportion of participants in each shedding duration group.

**Figure 2** shows the average SARS-CoV-2 viral load by day of viral shedding and by COVID-19 symptomatic status. In 73 persons who reported COVID-19 symptoms, the peak average viral load was observed on day 3 of viral shedding at a Ct value of 28.1. Among 24 asymptomatic persons, the average viral load peaked on day 3 of viral shedding (Ct= 36.4), then decreased before increasing again after day 7. The sample sizes for the average viral load calculations are presented in **Figure 2**.

**Figure 2.**
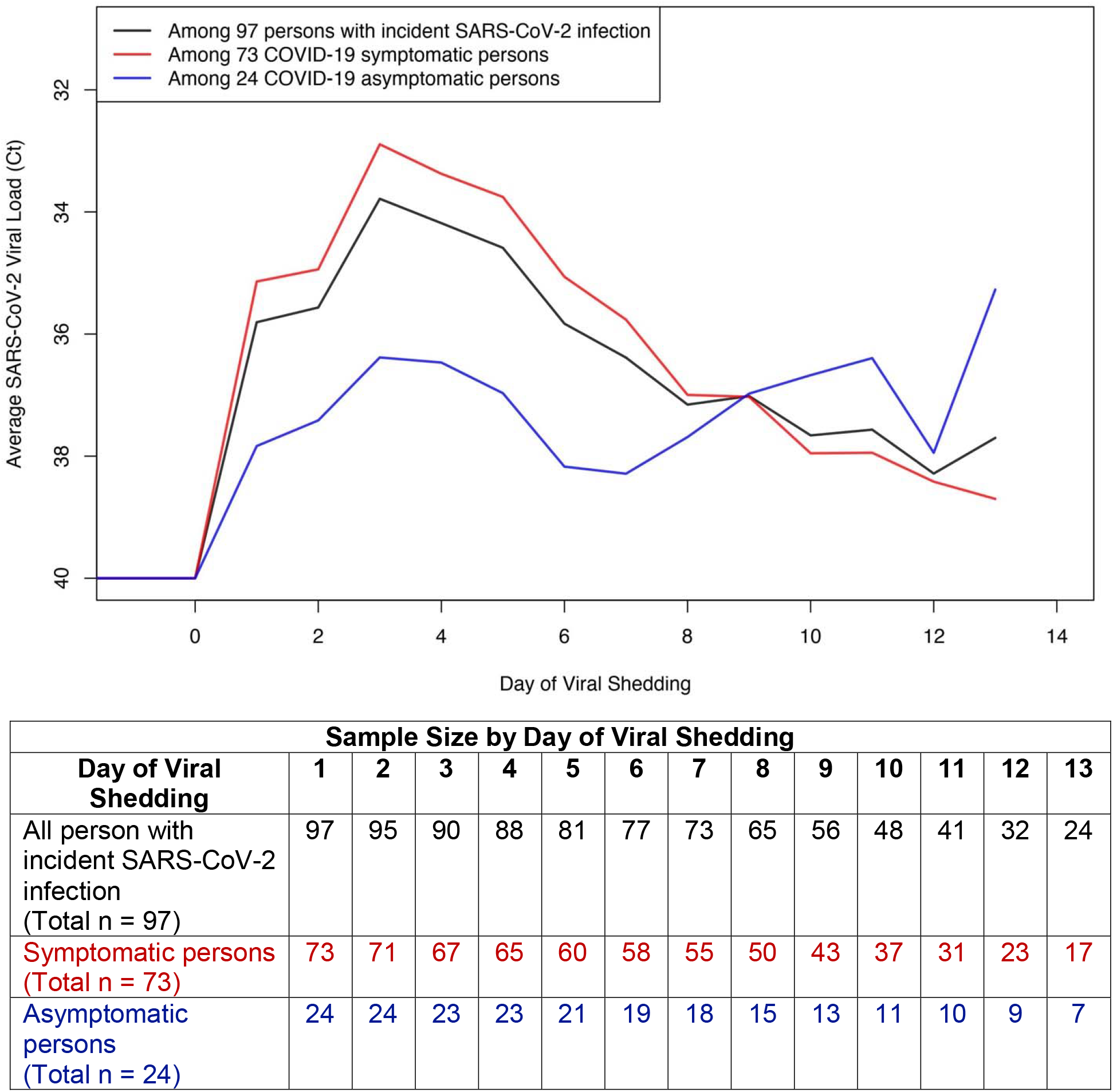
The average Severe Acute Respiratory Syndrome Coronavirus-2 (SARS-CoV-2) viral load cycle threshold (Ct) values by day of viral shedding among (a) the 97 persons with incident SARS-CoV-2 infection, (b) the 73 symptomatic persons, and (c) the 24 asymptomatic persons. The sample sizes for the average viral load calculations were presented in the table in the lower panel. The Centers for Disease Control and Prevention (CDC) clinical criteria were used to ascertain symptomatic Coronavirus disease 2019 (COVID-19 cases). Of note, less than 16 asymptomatic persons were used to calculate the average viral load after day 7 of viral shedding.

### Model-based viral shedding characteristics

**Figure 3** shows the model-based duration of shedding and peak viral load for participants with 2 or more days of observed viral shedding in the parent study (n=129).^28^ Fifty-three of the participants were SARS-CoV-2 negative at baseline, and 76 were SARS-CoV-2 positive at baseline. The estimated distribution of shedding duration was highly variable (median=11 days, IQR=7 – 15 days), with 32% lasting for more than 14-days (**Figure 3a**). The estimated peak viral load was variable based on the range of Ct (median Ct= 25, IQR Ct= 19 – 33), with an estimated 28% of the episodes peaking at Ct value <20. Viral load distribution was bimodal, with modes at Ct values of 36 – 37 and 19 – 20, respectively (**Figure 3b**).

**Figure 3(a).**
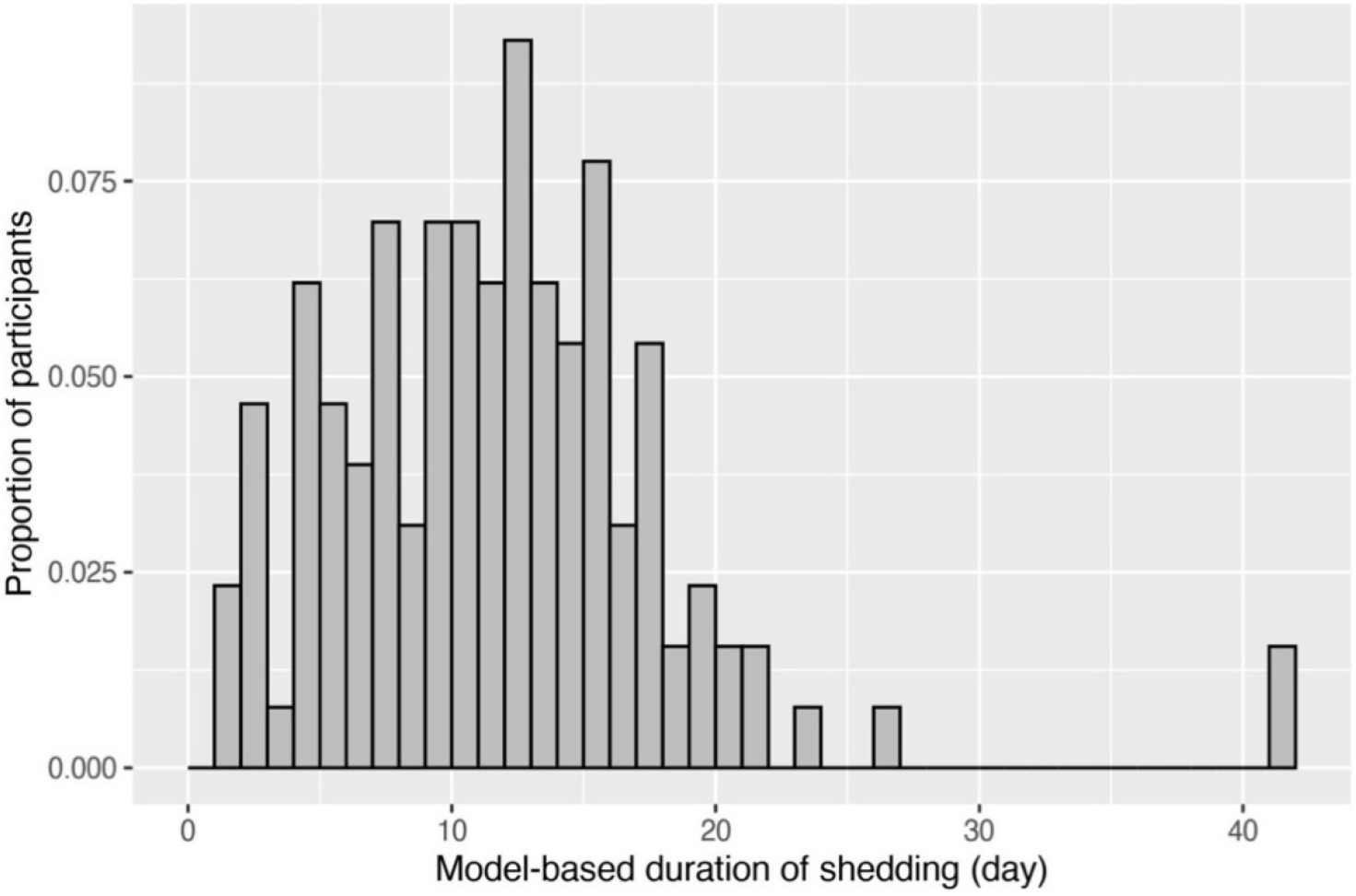
Histogram of the model-based duration of shedding among the 129 participants who had 2 or more days of observed Severe Acute Respiratory Syndrome Coronavirus-2 (SARS-CoV-2) viral shedding in the parent study.

**Figure 3(b).**
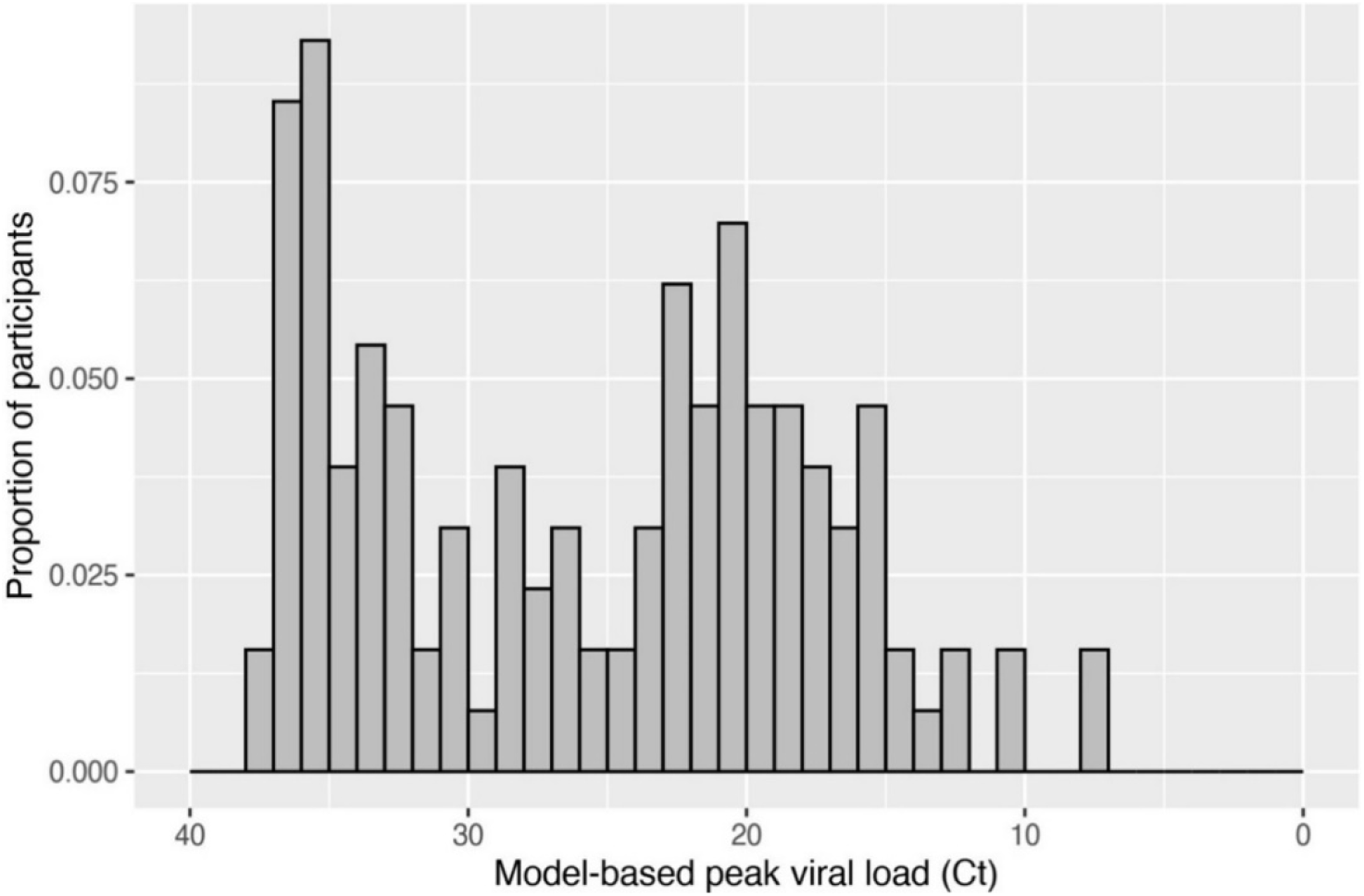
Histogram of the model-based peak viral load measured by cycle threshold (Ct) value among the 129 participants who had 2 or more days of observed Severe Acute Respiratory Syndrome Coronavirus-2 (SARS-CoV-2) viral shedding in the parent study.

**Figure 4** graphically summarizes the estimated stereotypical SARS-CoV-2 viral load trajectories for participants with different duration of shedding. The shedding episodes were characterized by a rapid upslope in viral shedding with a subsequent slower viral decay. The estimated correlation coefficient between peak Ct and shedding duration was -0.25 (95% credible interval, -0.33, -0.17), indicating that people who shed longer tended to have higher peak viral RNA. After shedding onset, the modeled median time to peak viral RNA was 1.4 days (IQR=0.9 – 1.9 days) compared to a median of 9.7 days (IQR=6.2 – 13.3 days) between the peak and the end of viral detection.

**Figure 4.**
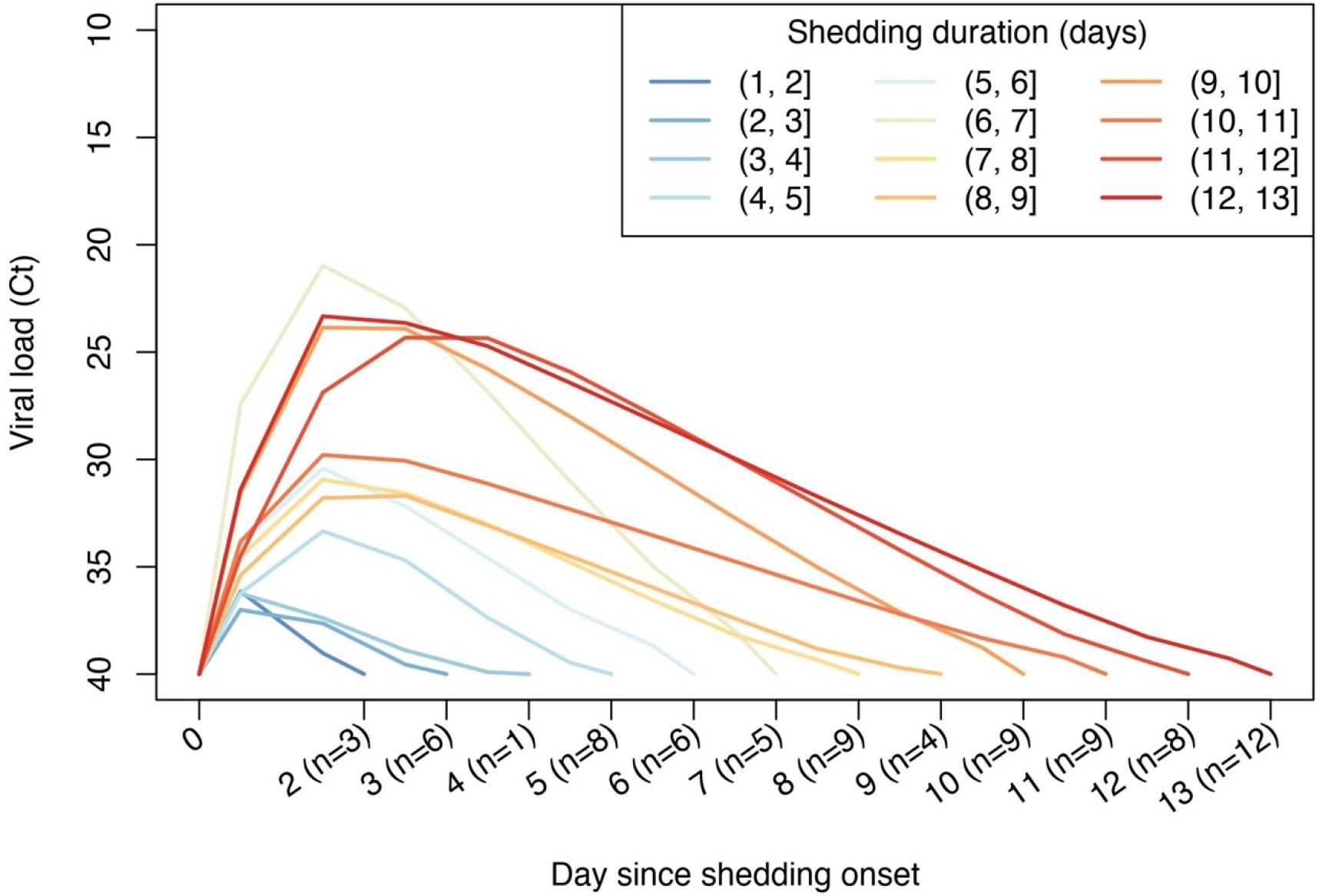
Model-based Severe Acute Respiratory Syndrome Coronavirus-2 (SARS-CoV-2) shedding characteristics. Summary curves of mean cycle threshold (Ct) value by day since shedding onset within each model-based duration group. The number of participants within each duration group was labeled on the x-axis.

## Discussion

Our study provides a detailed description of the G614 SARS-CoV-2 viral trajectories assessed prospectively by viral RNA detection in a diverse, COVID-19 vaccine-naïve, community-based outpatient cohort. The highest average viral load occurs quickly after viral shedding onset and at a higher magnitude in symptomatic than asymptomatic persons. The viral shedding episodes were notable for a sharp upswing to reach the peak viral load followed by a prolonged decay. The viral shedding duration varies widely and correlates with viral load peak. Our unique study complements and strengthens previously published observations and estimations of SARS-CoV-2 viral trajectories by providing a thorough evaluation with frequent molecular testing and detailed symptoms report collected daily after exposure to COVID-19 cases.

Our study confirms data from previous studies that documented a variable duration of viral RNA shedding ^19,35–37^ and expands those findings by providing early viral detection in asymptomatic persons with incident SARS-CoV-2 infection. One systematic review evaluating shedding data from 28 studies showed that the median duration of viral shedding was 18 days following onset of symptoms, with the longest period of viral RNA detection in the upper respiratory tract of 83 days.^38^ Of note, most studies (97%) included in the review were conducted among hospitalized patients and samples were obtained after COVID-19 symptom onset. However, in an out-of-hospital cohort of quarantined 303 patients with lab-confirmed SARS-CoV-2 infection in Korea, the median duration of viral RNA detection of 17-19 days was similar to that observed in hospitalized patients.^39^ Although asymptomatic and symptomatic patients included in the study received frequent SARS-CoV-2 testing from upper and lower respiratory tract samples (days 8, 9, 15, and 16 of isolation with an option to extend testing up to day 19), lack of early sampling after exposure to index cases could have missed early viral detection and accurate viral duration estimation. In our analysis of 97 adults with incident infection, the duration of SARS-CoV-2 G614 shedding in nasal swabs lasted from 1 to 13 days and was limited by the 14-day follow-up. According to our model informed by the observed data, the estimated shedding was 1 to 42 days. Of note, almost half of our participants had a single positive swab with lower quantities of SARS-CoV-2 detected compared to participants with multiple positive swabs. Although our study enrolled participants who reported exposure to at least one lab-confirmed case in their household or close contacts, precise exposures and timing of transmissions are challenging to validate. Some participants may have had asymptomatic infections prior to enrollment and we captured the end of their infection when viral detection could be intermittent.^20,23,40,41^ Single-day detection of SARS-CoV-2 RNA could also be related to the size of the viral inoculum allowing the innate immunity to clear the virus efficiently.^42^ Alternatively, immune priming by prior seasonal coronaviruses may contribute to limited infections.^43–45^ Furthermore, a few participants may have had pre-existing adaptive immunity from prior SARS-CoV-2 infection, as we did not document their antibody status at enrollment.^46–48^ As SARS-CoV-2 viral shedding duration is variable among individuals, frequent measurements early after exposure with sensitive viral detection assays are essential to quantify infection duration for fitting transmission models and assessment of therapeutic efficacy in trials using infection duration as endpoint.

Our estimate of a rapid viral peak and subsequent slower decline has been demonstrated in prior mathematical models of SARS-CoV-2 infection and other respiratory viruses using molecular detection such as RSV, HRV, H1N1, as well as chronic non-respiratory viral infections such as HSV.^37,47,49–53^ Our observation of lower viral load at the initiation of viral shedding with a peak viral load shortly after shedding onset suggests a rapid viral production following viral acquisition. The transition from the peak to the viral decay occurs quickly and the magnitude of the peak viral seems to dictate the duration of the shedding episode (intense and extended infection). The higher average viral load among persons who reported COVID-19 symptoms than in asymptomatic persons suggests that the copy number is predictive of clinical disease. Our results of higher viral load in symptomatic than asymptomatic persons agree with findings in a recent community-based evaluating RNA levels in self-collected nasal swabs from adults and children (n=550) in Washington State^54^ and a small (n=31) prospective study among patients hospitalized in China^55.^ However, others did not find associations between viral load and disease severity^36,56,57^ Emerging data indicate that SARS-CoV-2 viral load in respiratory samples is an important driver of SARS-CoV-2 transmission^58^; therefore, an accurate viral load measurement is an important parameter to develop mathematical models of viral spread. This is particularly important as preliminary findings indicate that new variants of concerns, such as the B.1.1.7 lineage, are associated with a significantly higher viral load than variants circulating early in the pandemic, which may explain their greater transmissibility. ^25,27^

Acute respiratory viral infections pose multiple challenges in the evaluation of therapeutics. The poor specificity of clinical symptoms for particular pathogens, variability in magnitude and duration of viral shedding, and the short window of viral replication affects the ability to evaluate candidate antivirals. Pragmatically, the SARS-CoV-2 viral dynamics also create challenges to devise scalable treatments that can be given early enough to affect the peak viral load and modify the natural history of the disease. With the widespread use of the COVID-19 vaccines, placebo-controlled clinical trials for evaluation of new preventive interventions will become more challenging to design and implement. These data provide accurate assessment of viral trajectories absent effective intervention and can form the basis for the design of future trials or essential historical controls.

## Limitations

Our study has several limitations. Self-sampling technique or specimen degradation could yield false-negative results. However, unsupervised home mid-turbinate swab collection for SARS-CoV-2 detection has been demonstrated comparable to clinician-collected samples and samples are stable at ambient temperature for up to 9 days.^59,60^ The universal RNase P DNA detection confirmed that participants were successful in the collection of a valid sample, and the high incidence of SARS-CoV-2 based on self-collected swabs suggests that self-sampling was adequate. Our analysis is limited to the 14-day observation period which underestimates shedding duration due to right censoring. Furthermore, despite collecting frequent swabs early after exposure to persons with confirmed SARS-CoV-2 infection, daily swabbing could have missed the peak viral load in those with shorter shedding duration. Our viral shedding model assumed that viral load followed a piece-wise linear trend, whereas the true trajectories might be more curved. Due to incomplete observed trajectories and missing data, the model-based trajectories for many participants were associated with large limitations.

## Conclusions

Our study of the early SARS-CoV-2 G614 variant infections suggests that the viral replication peaks rapidly followed by a slow decline suggesting that a period for effective antiviral intervention is very narrow. Repeated testing for SARS-CoV-2 could substantially increase detection rates among exposed close contacts. Our longitudinal evaluation of the SARS-CoV-2 G614 variant before implementing COVID-19 vaccines may serve as a reference for comparison of emergent viral lineages to help inform clinical trial design and public health strategies to contain the spread of the virus and ongoing therapeutic development for SARS-CoV-2.

## Data Availability

Relevant data are available from the corresponding author on request.

