## Supplemental materials for "Trajectory of viral load in a prospective population-based cohort with incident SARS-CoV-2 G614 infection"

***Figure S3. Model-based individual-level SARS-CoV-2 viral load trajectories……………………….…..7***

### ***Table S1. Summary of SARS-CoV-2 lineages………………………………………………………………………….13***

### Supplemental methods – Piece-wise linear mixed-effect model for viral shedding.

Since we only collected nasal swabs from each participant for 14 consecutive days, many (108 out of 180) participants who shed during the follow-up period had censored shedding, i.e., their first and/or last collected swabs tested positive for Severe Acute Respiratory Syndrome Coronavirus-2 (SARS-CoV-2). We fit a Bayesian piece-wise linear mixed-effect model to estimate the participants’ total duration of viral shedding and the peak viral load.

**Data**

We used data from 129 participants who had 2 or more days of observed viral shedding during the first 14 days of follow-up in the parent study. Fifty-three of the participants were SARS-CoV-2 negative at baseline (i.e., incident SARS-CoV-2 infection), and 76 were SARS-CoV-2 positive at baseline. The participants who only had 1 observed day of shedding were excluded from this model because we were unable to ascertain whether the single positive swab represented the tail of an infection acquired before study entry, a re-infection, or a false-positive result. The participants who were SARS-CoV-2 positive at baseline were included to enrich the observed viral load trajectory data, especially for estimating the rate of decay of viral load.

**Model description**

We fitted a piece-wise linear mixed-effect model to estimate the peak viral load measured in cycle threshold (Ct) value, the time from shedding onset to peak viral load, and the time from peak to viral clearance. We assumed that the viral load trajectories follow a trend that consists of a proliferation phase with linear growth of viral load on the Ct scale, followed by a clearance phase with linear decay of viral load on the Ct scale. This corresponds to exponential growth and delay in viral RNA concentration in the respective phases. This idealized trajectory is depicted in **Figure S1** of this supplement and represented by the following equations:

$$E\left[ Ct\left( s \right) \right]=40-\mu_{y}\left( s \right)$$

$$\mu_{y\left( s \right)}=v_{p}\times I\left[ -w_{a}\leq s\leq w_{b} \right]+\frac{v_{p}}{w_{a}}\times s\times I\left[ -w_{a}\leq s\leq w_{b} \right]-\frac{v_{p}}{w_{b}}\times s\times I\left[ -w_{a}\leq s\leq w_{b} \right]$$

Here, $I\left( \cdot\right)$ represents an indicator function. $s$ represents time since the peak viral load of the idealized viral trajectory, so that $s = 0$ at the peak of the trajectory. $E\left[ Ct\left( s \right) \right]$ represents the expected value of viral load in Ct at time $s.$ $\mu_{y}\left( s \right)$ represents the difference between the level of detection (LOD, equals 40) and the expected viral load at time $s$. $v_{p}$ represents the absolute difference between LOD and the peak viral load in Ct. $w_{a}$represents time in days between shedding onset and peak viral load, and $w_{b}$represents time in days between peak viral load and viral clearance. $t_{p}$ represents the time difference between the latent peak viral load and the observed peak viral load.

We assumed that the difference between the observed viral load and the LOD, $y\left( s \right)$, has the following distribution:

$$y\left( s \right)\sim\text{Normal}\left( \mu_{y}\left( s \right),\sigma_{y}\left( s \right) \right)$$

$$\sigma_{y}\left( s \right)=\sigma_{yy}\times I\left[ -w_{a}\leq s\leq w_{b} \right]+0.05\times\left( 1-I\left[ -w_{a}\leq s\leq w_{b} \right] \right)$$

This model assumes that the observed Ct values are normally distributed around the expected trajectory with standard error $\sigma_{yy}$ during the viral shedding, and with standard error $0.05$ before and after viral shedding to allow for deviation from LOD due to potential errors from various sources such as misplaced swabs.

We used random effects to capture individual-level variations from the population-level means ($\mu$) and specified the following distributions for the random effects:

$$t_{p,i}\sim_{i.i.d.}\text{Normal}\left( \mu_{tp},\sigma_{tp} \right)$$

$$\log\left( \left[ v_{p,i},w_{a,i},w_{b,i} \right] \right)\sim\text{Multivariate Normal}\left( \mu_{log},\Sigma_{log} \right)$$

Here, $\mu_{log}$ is a vector of length 3 and $\Sigma_{log}$ is a 3 by 3 variance-covariance matrix.

We used a Markov Chain Monte Carlo (MCMC) fitting procedure implemented in JAGS (version 4.3.0)^1^ and R (version 4.0.3)^2^ to estimate the individual-level and population-level parameters. We incorporated information from previous work^3^ by specifying the following prior distributions for the population-level means:

$$\mu_{tp}\sim\text{Normal}\left( 0,1 \right)$$

$$\mu_{log}\left[ 1 \right]\sim\text{Normal}\left( \log\left( 18 \right),2 \right), \text{truncated}\left[ -\infty,\log\left( 40 \right) \right]$$

$$\mu_{log}\left[ 2 \right]\sim\text{Normal}\left( \log\left( 3 \right),1 \right)$$

$$\mu_{log}\left[ 3 \right]\sim\text{Normal}\left( \log\left( 9 \right),1 \right)$$

We used weakly-informative priors for the rest of the parameters:

$$\sigma_{tp}\sim\text{Cauchy}\left( 0,1 \right)\text{, truncated}\left[ 0,\infty\right]$$

$$\sigma_{yy}\sim\text{Cauchy}\left( 0,1 \right)\text{, truncated}\left[ 0,\infty\right]$$

$$\Sigma_{log}\sim\text{Inverse Wishart (}I_{3\times3}\text{ , 4)}$$

We ran an MCMC chain for 10,000 iterations, with the first 5,000 iterations discarded as burn-in. Using a thinning interval of 5, we used 1,000 of the second 5,000 iterations for inference.

**Summary results**

The posterior distributions of key population-level parameters are shown in **Figure S2**. Specifically, we plotted the histograms of 1,000 posterior samples of the population-level mean (a) peak viral load, 40 - $v_{p}$, (b) time from shedding onset to peak viral load, $w_{a}$,(c) time from peak viral load to viral clearance, $w_{b}$, and (d) total duration of shedding, ${w_{a}+w}_{b}$.

In **Figure S3**, we showed the best-fit viral load trajectories with 90% credible intervals for each of the 129 participants who had 2 or more days of observed viral shedding during the first 14 days of follow-up. Care must be taken when considering the model-based duration of shedding and peak viral load. For example, 2 participants (PID A66 and PID A93) had model-based shedding duration of more than 40 days; however, examining the estimated trajectories with uncertainty intervals revealed that the estimates were heavily influenced by a few data points near the end of follow-up period, and the uncertainty intervals around the viral clearance times were very wide.

### **Figure S1**. A theoretical SARS-CoV-2 viral load trend (blue line) overlaying the observed viral load for a participant in the study.

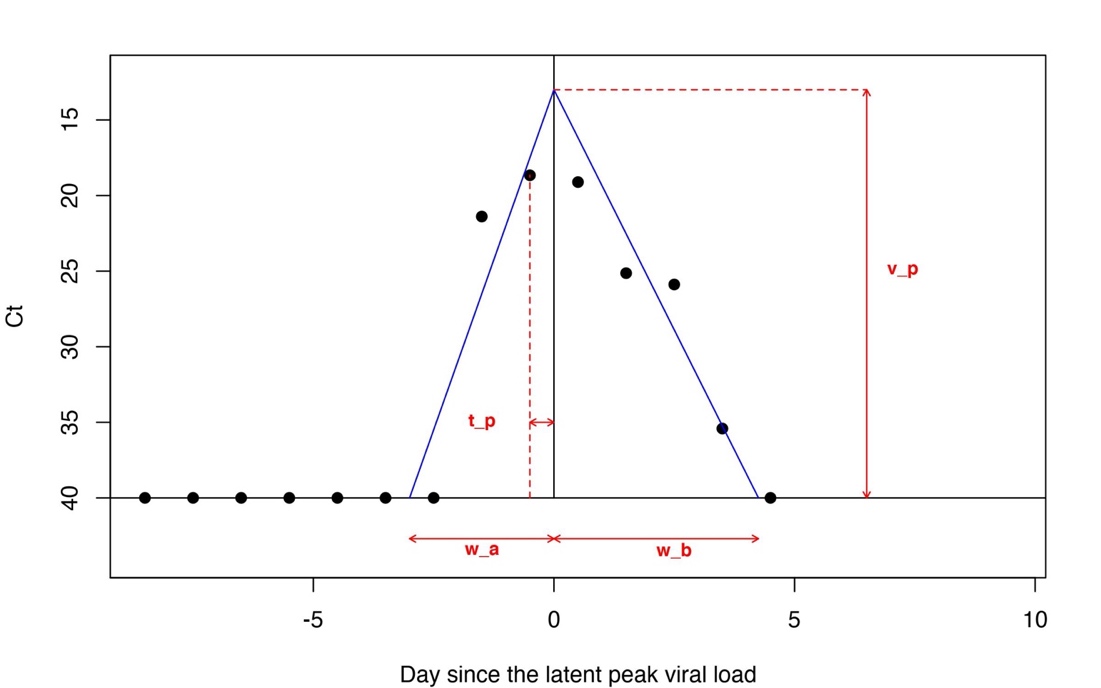

### **Figure S2**. Posterior distributions of population-level mean peak viral load and duration of viral shedding stages.

(a) Histogram of 1,000 samples from the posterior distribution of the population-level mean peak viral load (Ct).

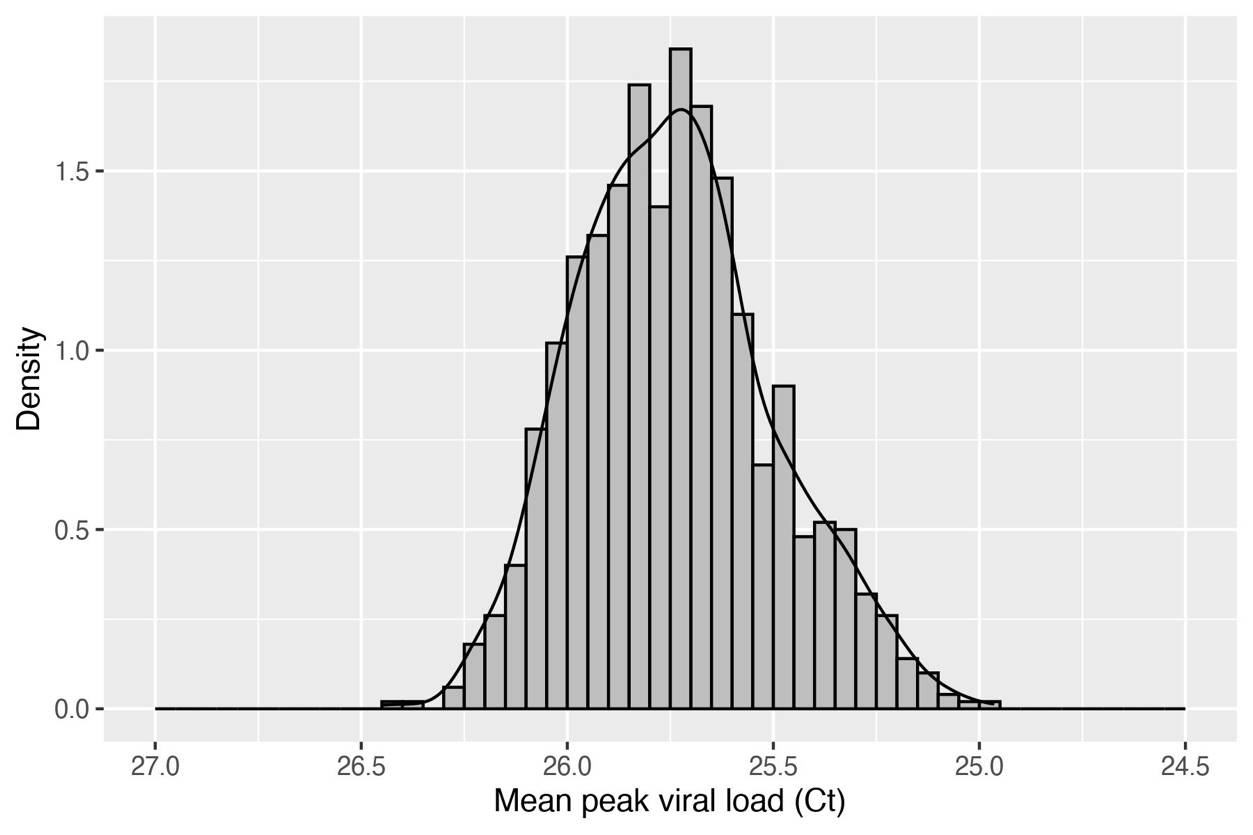

(b) Histogram of 1,000 samples from the posterior distribution of the population-level mean time from shedding onset to peak viral load (measured in days).

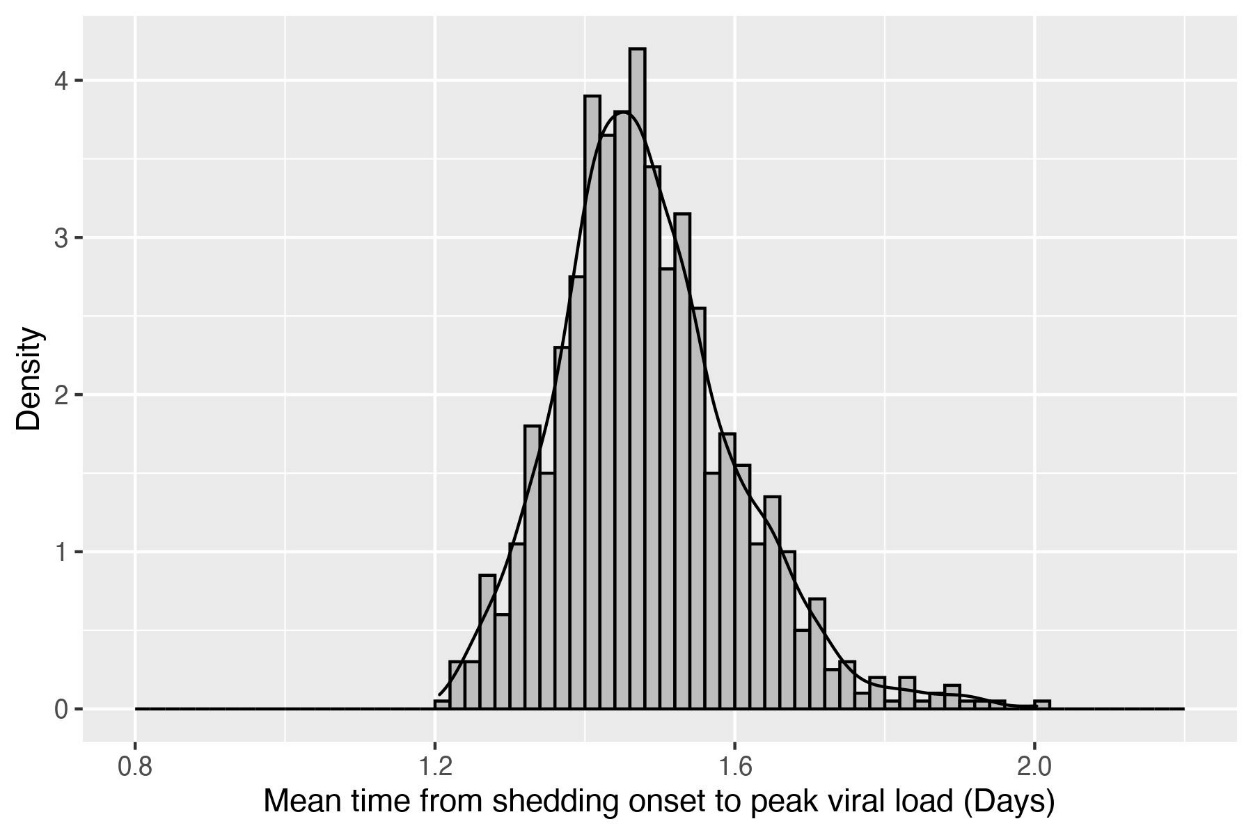

(c) Histogram of 1,000 samples from the posterior distribution of the population-level mean time from peak viral load to viral clearance (measured in days).

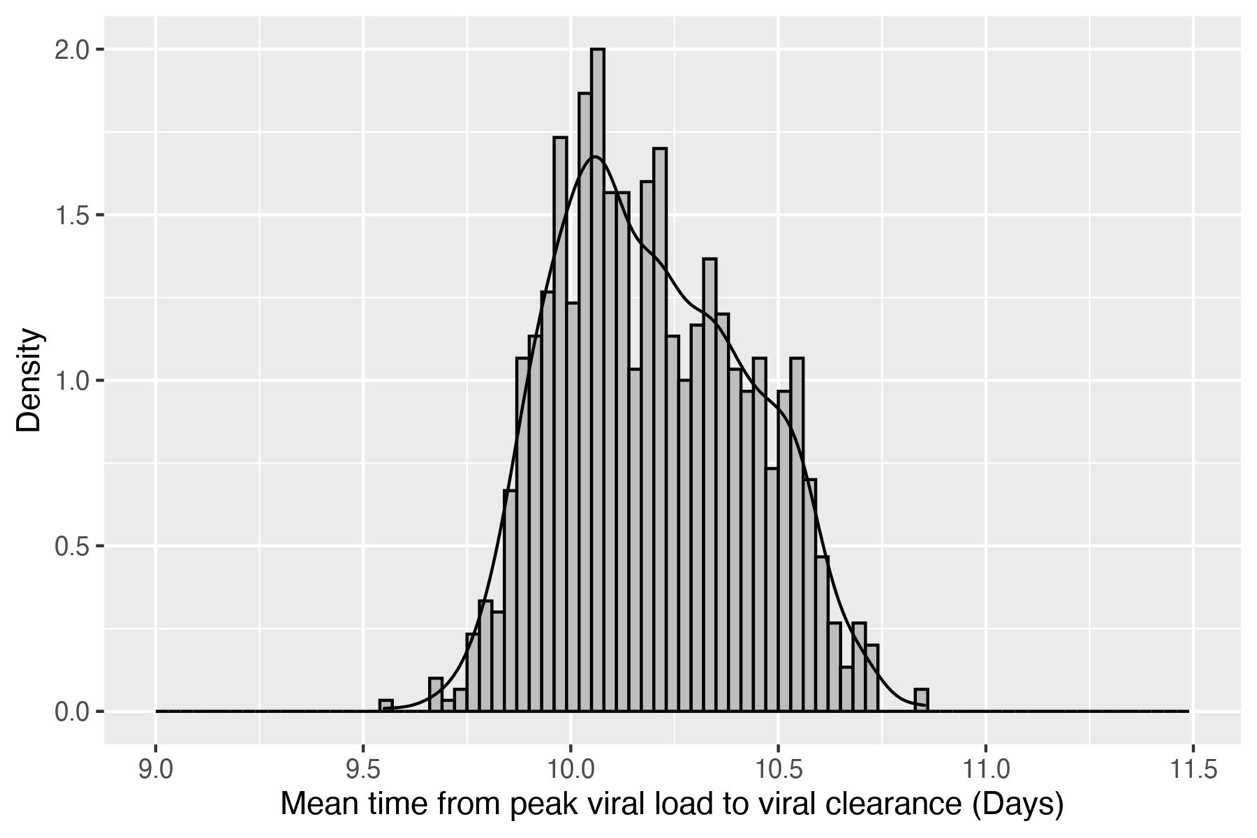

(d) Histogram of 1,000 samples from the posterior distribution of the population-level mean duration of shedding (measured in days).

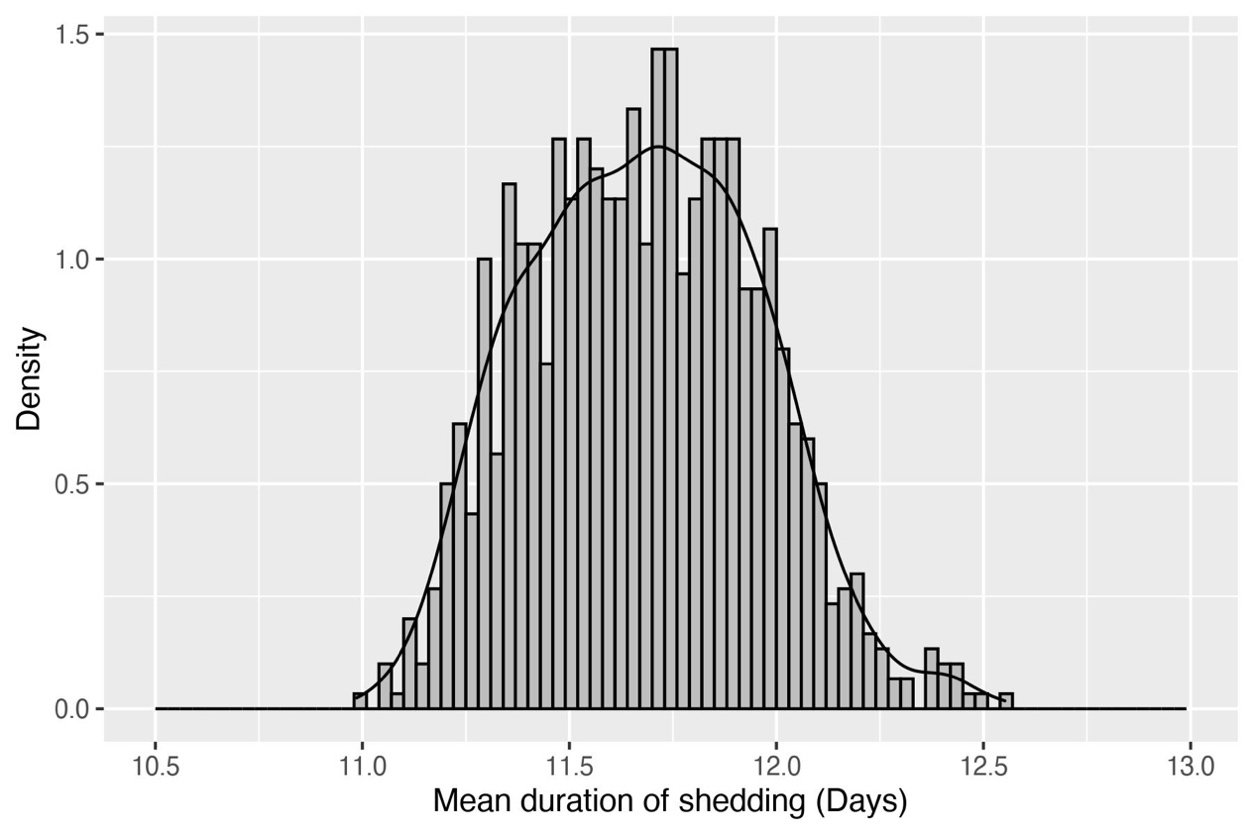

**Figure S3.** Model-based individual-level Severe Acute Respiratory Syndrome Coronavirus-2 (SARS-CoV-2) viral load trajectories for each participant who shed for 2 or more days during the first 14 days of follow-up in the parent study, centered at the day of the observed peak viral load. The red lines and ribbons were the posterior mean and the 90% credible interval of SARS-CoV-2 viral shedding, and the black dots were the observed SARS-CoV-2 viral cycle threshold (Ct) values from Polymerase Chain Reaction (PCR) tests.

a. Among 53 participants with a negative swab for SARS-CoV-2 at baseline.

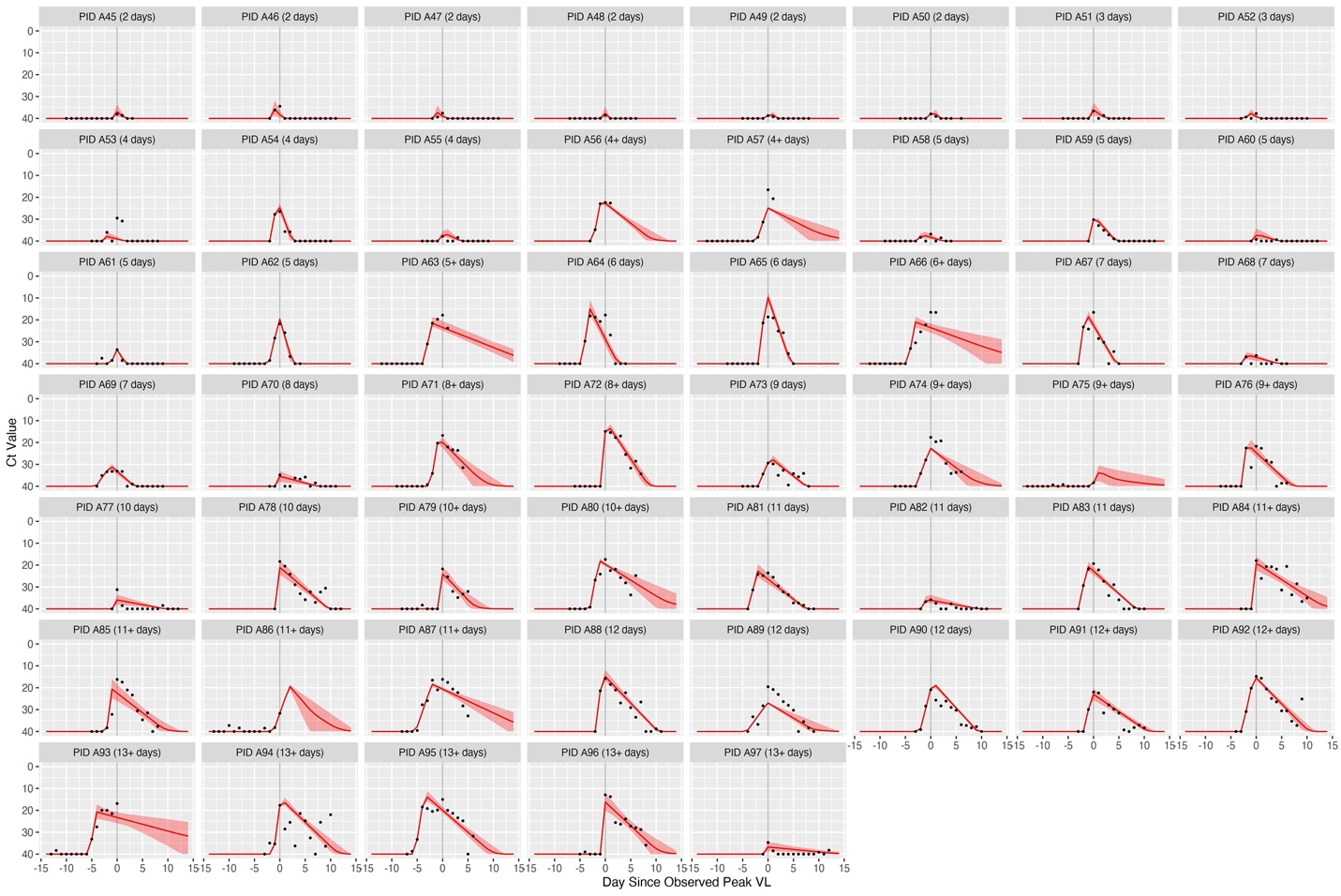

b. Among 76 participants with a positive swab for SARS-CoV-2 at baseline.

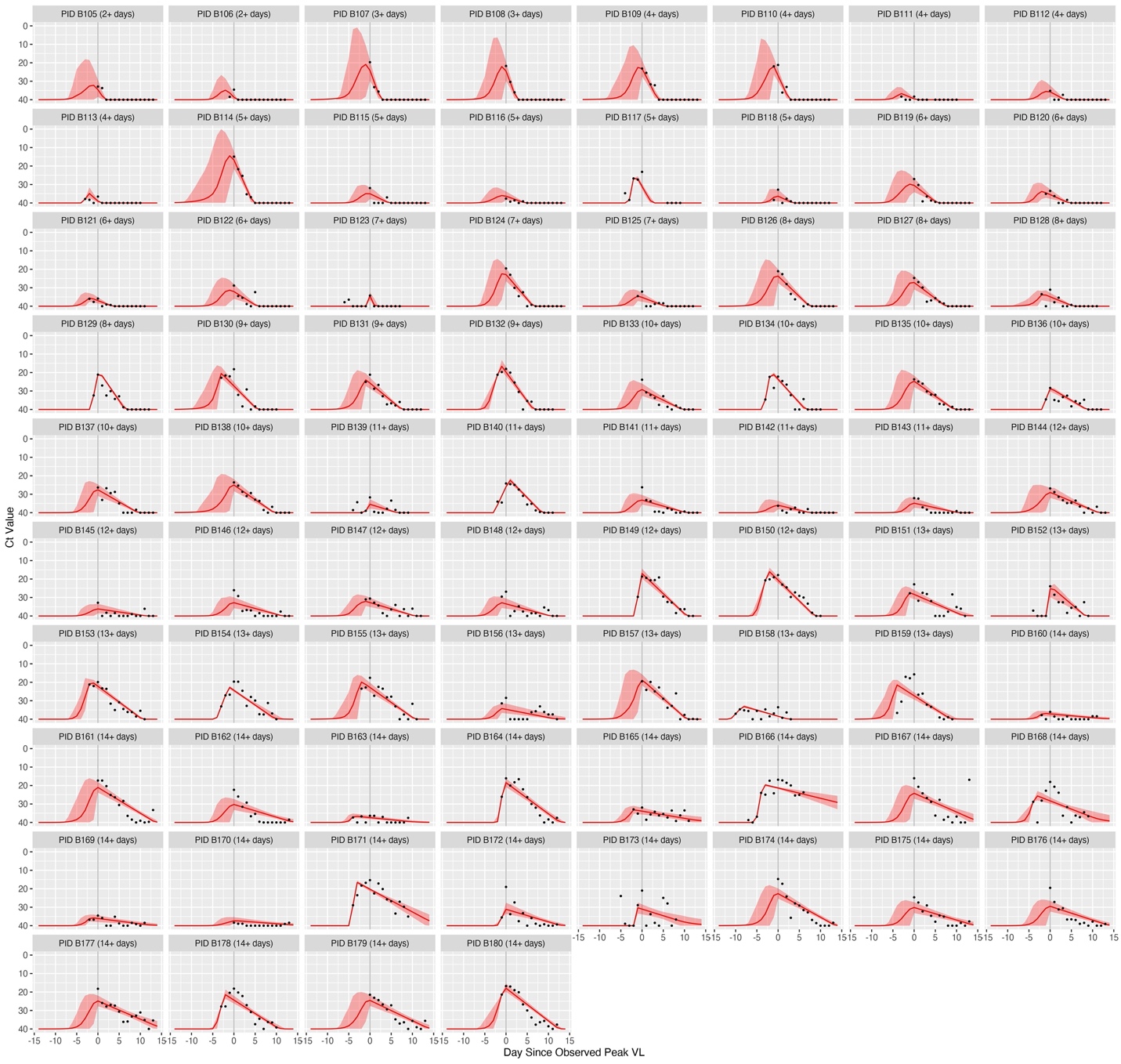

**Figure S4**. The observed viral load trajectories for 97 participants with incident Severe Acute Respiratory Syndrome Coronavirus-2 (SARS-CoV-2) infection. The observed viral shedding duration for each participant was defined as the number of days from the first positive self-collected mid-turbinate swab for SARS-CoV-2 until the last positive swab. Censoring was indicated if a participant had a positive swab on the last day of swab collection. For a given day, a participant’s viral load was calculated as the average of N1 and N2 Ct values from the Reverse Transcription Polymerase Chain Reaction (RT-PCR) test. The cycle threshold (Ct) value assigned to the negative samples was 40. Each “PID” represents a participant and the number of days in parentheses indicates the observed duration of SARS-CoV-2 shedding, with “N+” days indicating censoring. Each dot represents 1 self-collected mid-turbinate swab and their corresponded cycle threshold value is represented on the y-axis.

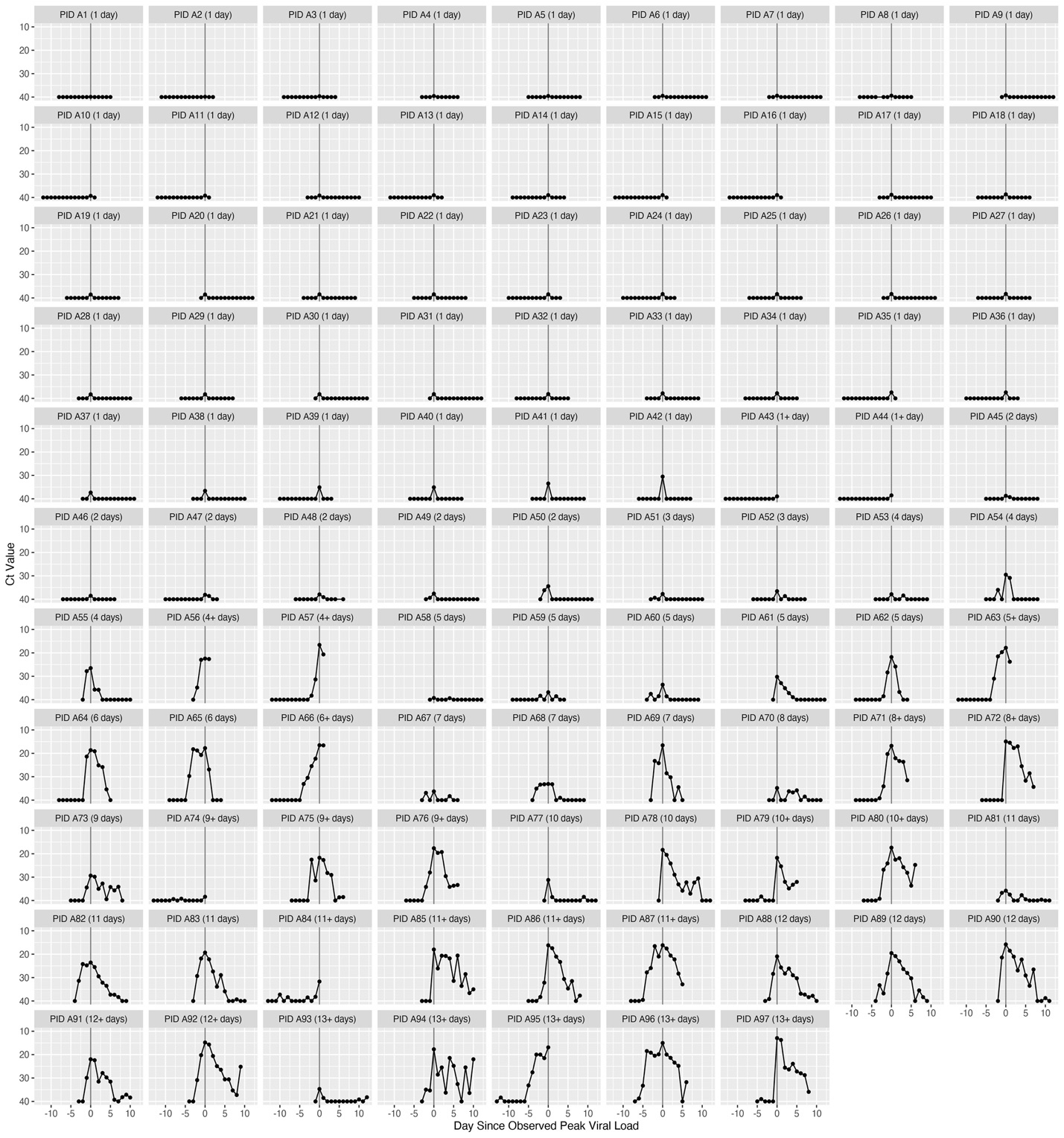

### **Figure S5.** Spaghetti plots for the observed Severe Acute Respiratory Syndrome Coronavirus-2 (SARS-CoV-2) viral load trajectories among 97 participants with incident SARS-CoV-2 infection, centered at day of the observed peak viral load (measured as cycle threshold, Ct) and grouped by the duration of shedding. The blue lines and the grey ribbons were the locally weighted scatterplot smoothing (LOWESS) curves and the 95% confidence intervals.

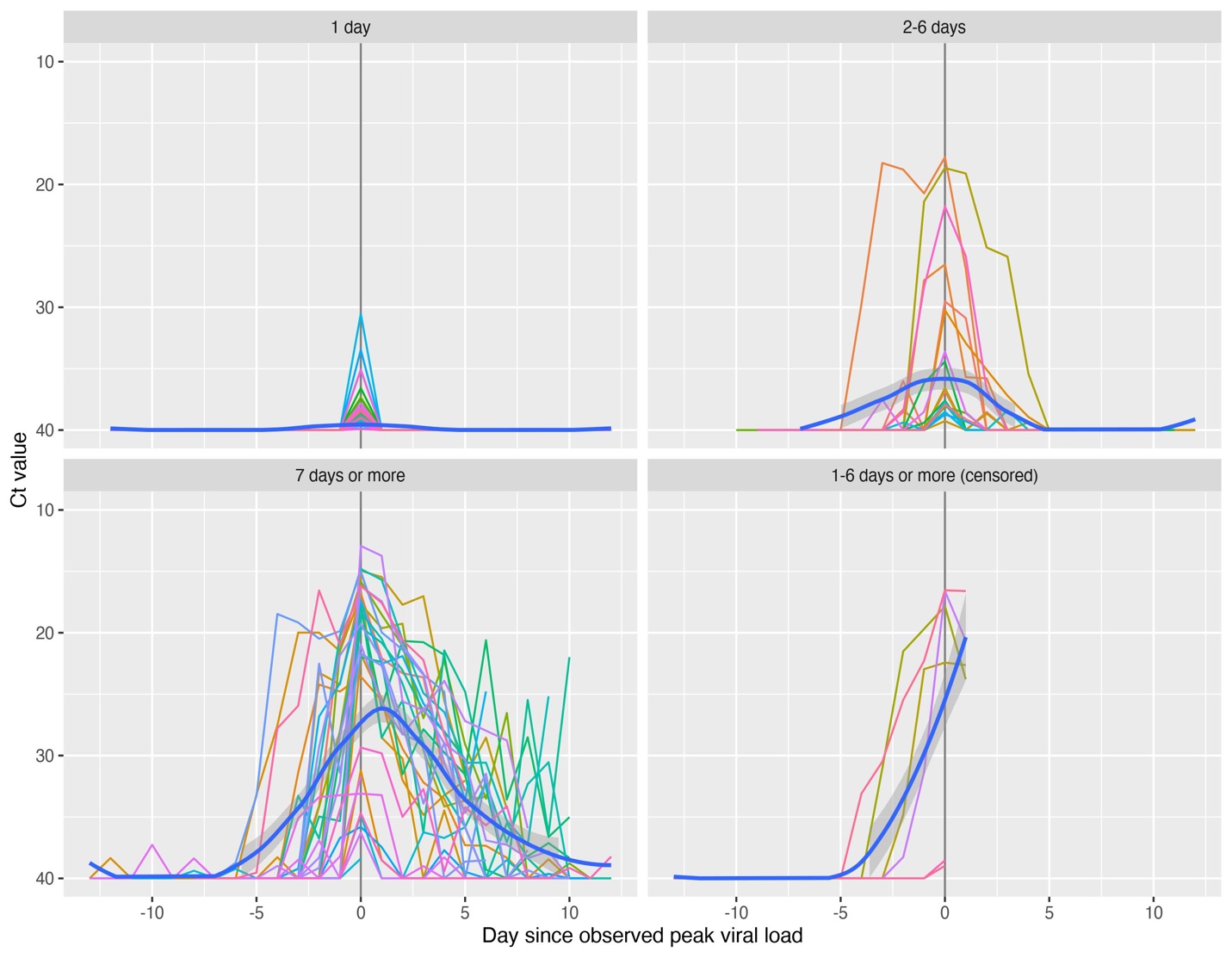

### **Figure S6.** Distribution of the RNAse P cycle threshold (Ct) values.

(a) Distribution of the RNAse P Ct values, grouped by the  Severe Acute Respiratory Syndrome Coronavirus-2 (SARS-CoV-2) PCR status. Each dot represents one observed RNAse P Ct value obtained among all study participants in the parent study.^4^ The horizontal line segments are the median RNAse P Ct values categorized by SARS-CoV-2 detection.

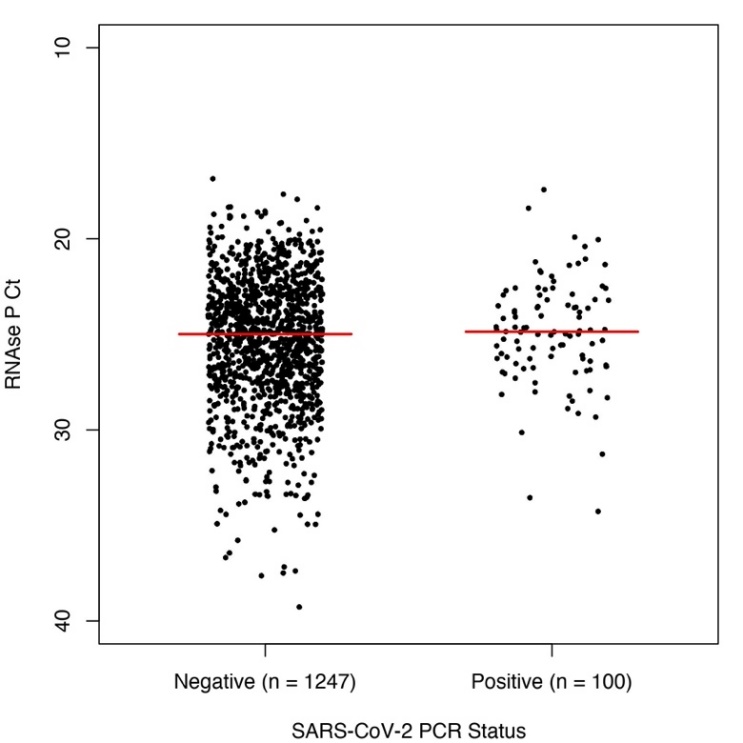

(b) Scatterplot of the RNAse P Ct values against the SARS-CoV-2 viral load Ct values among all positive SARS-CoV-2 samples that were tested for RNAse P in the parent study (n = 100), with the locally weighted scatterplot smoothing (LOWESS) curve and the 95% confidence interval.

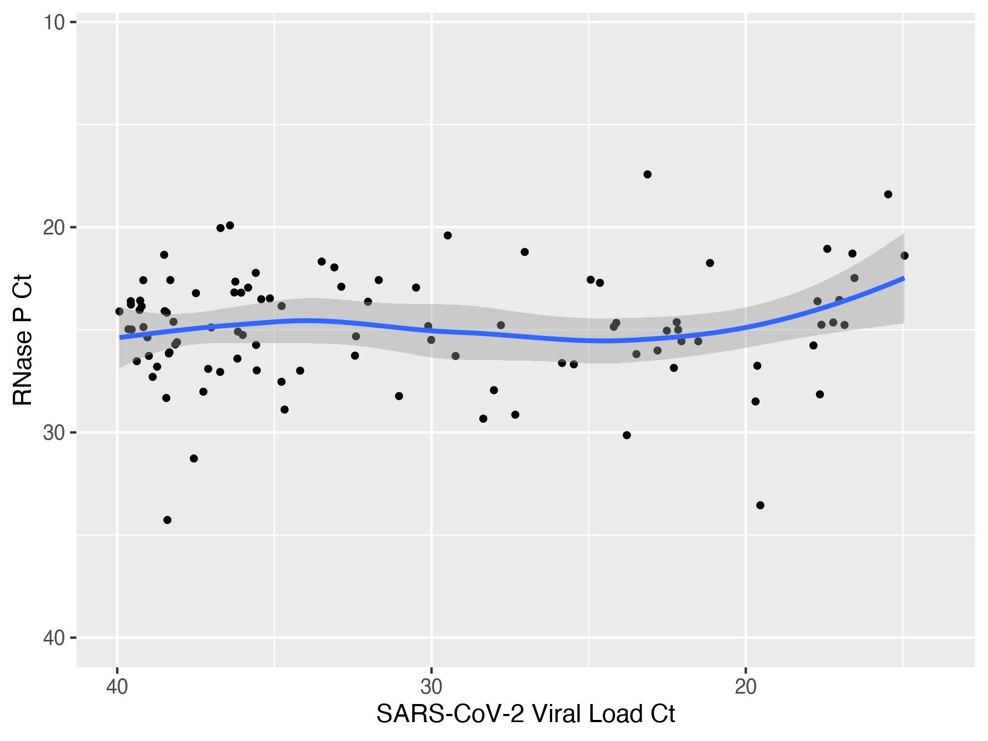

| **Table S1.** Summary of SARS-CoV-2 lineages observed in the study. A total of 215 samples with Ct<34, corresponding to 42 participants were sequenced. | |
| --- | --- |
| **Lineage** | **Number of samples** |
| B.1 | 102 |
| B.1.369 | 24 |
| B.1.1 | 14 |
| B.1.371 | 14 |
| B.1.594 | 14 |
| B.1.1.291 | 12 |
| B.1.509 | 10 |
| B.1.516 | 9 |
| B.1.1.186 | 5 |
| B.1.240 | 5 |
| B.1.111 | 2 |
| B.1.1.29 | 1 |
| B.1.36.36 | 1 |
| B.1.400 | 1 |
| B.1.576 | 1 |
